# Analysis of Crowdsourced Metformin Tablets from Individuals Reveals Widespread Contamination with N-Nitrosodimethylamine (NDMA) and N,N-Dimethylformamide (DMF) in the United States

**DOI:** 10.1101/2020.05.22.20110635

**Authors:** Qian Wu, Evgenia Kvitko, Amber Hudspeth, Shannon Williams, Ryan C. Costantino, Kaury Kucera, David Light

## Abstract

Reports of metformin drug products contaminated with unacceptable levels of the probable human carcinogen N-Nitrosodimethylamine (NDMA) prompted a national sampling of post-market metformin drug products in early 2020. To broadly sample the United States market and minimize supply chain bias, metformin medication samples were crowdsourced directly from individuals across many states. 155 samples were received, and liquid chromatography-high resolution mass spectrometry tests for a panel of nitrosamines and N,N-Dimethylformamide (DMF) revealed significant levels of NDMA and DMF that relate to formulation. 49% of all medication samples contained detectable levels of NDMA and, when scaled to maximum daily tablet dose, 16% of all medication samples contained NDMA levels exceeding the United States Food and Drug Administration acceptable daily intake (ADI) limit. The highest NDMA detection from the tested samples was 748 ng per 500 mg tablet, which, when scaled to a common 2000 mg per day dosage regimen, is 31 times the ADI limit. The presence of N,N-Dimethylformamide (DMF) across 74% of the sampled metformin products is concerning given its same carcinogenicity categorization as NDMA and proposed role in formation of NDMA. Results underscore the need for continued surveillance of product quality, recalls of tainted medications, and investigation of metformin manufacturing practices.

## Introduction

Metformin is one of the frontline medications used to treat type 2 diabetes and prediabetes. It is commonly prescribed when a patient exhibits consistently high blood sugar levels due to the inability of insulin to effectively transport sugar into somatic cells^1^ and can be taken for extended periods of time as a maintenance medication. Metformin is prescribed as a combination medication, solution, and suspension, but is commonly taken in 500 mg, 850 mg, and 1000 mg immediate release (IR) and 500 mg, 750 mg, and 1000 mg extended release (ER) tablets one or more times per day with a recommended maximum daily intake of 2,550 mg IR and 2,000 mg for ER.^2, 3^ Approximately 85 million prescriptions were written for metformin in the United States in 2019.^4^ Some research has likely led people to use metformin off-label as an alternative treatment for prediabetes, polycystic ovarian syndrome, fatty liver disease, weight loss, dementia prevention,^5^ thyroid disorders, and cancer prevention.^6^ Metformin is the only oral medication approved for diabetes treatment in children and has also been used to treat obesity in children and adolescents.^7^

Aspects of the metformin manufacturing process could lead to contamination with genotoxic compounds with carcinogenicity. One such contaminant, N-Nitrosodimethylamine (NDMA), a nitrosamine compound, is a probable human carcinogen according to the World Health Organization (WHO) and the International Agency for Research on Cancer (IARC).^8, 9^ Nitrosamines formed from reactive pharmaceutical products, or as by-products in pharmaceutical synthesis that contaminate drug products, have been extensively studied due to health risks since the 1970s.^10, 11^ Nitrosamine compound formation and contamination in drug products is currently a global concern. These probable human carcinogens may form when manufacturers reuse solvents that are neutralized with nitrite containing compounds in later steps of drug or medication processing. One common example is the use of the organic solvent, N,N-dimethylformamide (DMF) quenched with sodium nitrite, which has been proposed as the major cause of some NDMA contamination.^12^ On the other hand, DMF itself is considered to be a probable human carcinogen, thus making its detection in valsartan and metformin drug products of great concern.^13, 14^

The FDA currently lists the maximum acceptable daily intake (ADI) of NDMA at 96 ng^15, 16^ in accordance with the WHO’s interim guidelines, which were set in place in January 2020 to allow manufacturers to adjust procedures to minimize possible nitrosamine contamination.^12^ In 2021,the United States Pharmacopeia released methods for determining and limiting the nitrosamine content of medications^17^ following the 2018 FDA guidance document for industry.^15^

Instances of nitrosamine contamination were detected in mid-2018 in angiotensin II receptor blocker (ARB) drugs and the FDA announced voluntary recalls of three batches of valsartan.^18^ The initial recall was expanded to encompass several repackagers and two other ARB medications, irbesartan, and losartan as testing indicated levels of NDMA, N-Nitrosodiethlyamine (NDEA), and N-nitroso-N-methyl-4-aminobutyric acid (NMBA) exceeding the FDA’s acceptable limits. As of January 2022 in the U.S. market, a total of 1246 potentially contaminated ARB products have been affected by recalls.^19^

Reports of nitrosamine contamination have since grown to include heartburn and diabetes medications like ranitidine^20^ and metformin.^21^ Singapore was the first to take action against NDMA contamination in metformin, testing forty-six batches of the medication available in that country. Of those products, three were found to contain unacceptable levels of NDMA and were recalled in early December 2019.^22^ Switzerland^23^ and Canada^24^ soon launched their own investigations following Singapore’s recall. In Switzerland, Streuli Pharma recalled twenty batches of metformin products^25^ in mid-December 2019. Health Canada announced its initial recalls in February 2020, pulling ten batches with excessive levels of NDMA and four more with levels approaching the acceptable limit. One month later, an additional twenty-six lots were recalled in Canada as a precautionary measure.^26^

On February 3, 2020, the FDA released lab results for products from seven companies and sixteen lots of metformin showing concentrations of NDMA that would result in exposure below the ADI limit.^21^ Notably, the FDA may acquire drug or medication samples for analysis through voluntary submission by, or seizure from, specific labeling or manufacturing companies, which can generate sampling bias.^27, 28^ On March 2, 2020, an FDA citizen petition was filed by Valisure, LLC which contained lab results for twenty-two companies and thirty-eight lots of metformin showing widespread contamination with NDMA.^29^ Valisure acquired samples through its licensed pharmacy that purchased the metformin from United States-based distributors. Although this approach may reduce selection bias, it is still limited to the available products from a few specific distributors in a short period of time. On May 28, 2020, the FDA announced the finding of NDMA levels exceeding ADI limits in eight lots of metformin ER products made by five firms and requested voluntary recalls.^30^

Regulations for DMF are much less stringent compared to NDMA. Although potential carcinogenicity places both chemicals in the same Group 2A category, the current FDA permitted daily exposure (PDE) limit for DMF in pharmaceuticals is 8,800,000 ng.^31^ Valisure submitted a citizens petition to FDA in June of 2019 requesting stricter limits to be placed on this compound after high levels of contamination were found in valsartan products.^13^ Due to DMF’s potential contribution to the creation of NDMA and other carcinogens, a 2020 report from the European Medicines Agency recommends pharmaceutical manufactures not use DMF unless no alternate solvent is possible.^32^ While the synthetic steps used to prepare metformin are proprietary, there are patents for the synthesis of Metformin that employ DMF as a solvent and DMF has been observed recently as a residual solvent in metformin samples by other researchers.^33, 34^ Indeed, an early analysis of the dataset presented here overestimated NDMA levels in some metformin drug products because the original chromatography used did not fully separate DMF and NDMA. The concentration of DMF was, in some cases, so high that a rare nitrogen-15 isotope of DMF, with similar ion mass to NDMA, inflated NDMA quantification in some samples.^35^ Collaborative evolution of methods has enabled separation of NDMA and DMF and re-evaluation of the same dataset to reveal concerningly high DMF levels.

To minimize sampling bias as much as possible, Valisure crowdsourced samples of metformin directly from individuals in many states within the United States between mid-March to early August 2020. The availability of testing was publicized through social media channels including Facebook and Twitter, consumer advocate networks including The People’s Pharmacy, and other means. The testing program was open to people in all states except Massachusetts and Virginia due to restrictions, and individuals whose medications were paid for by government funded healthcare programs.

## Materials and Methods

In 2020, FDA published a series of methodologies on detection of nitrosamine compounds in metformin drug products using liquid chromatography high resolution mass spectrometry (LC-HRMS).^35-37^ Following the method testing principle, a modified method was validated in Valisure’s ISO 17025 accredited analytical laboratory to achieve a lower limit of quantification (LOQ), and to chromatographically separate NDMA and its interference compound ^15^N-DMF, thus to simultaneously detect NDMA and DMF in one LC-HRMS method. The method limits the potential for analytical artifacts originating from different inactive ingredients present in different formulations of metformin tablets made by different companies (this method is provided in detail on a pre-print server for public access).^38^

### Metformin Drug Product Sampling

Metformin drug product samples were crowdsourced from individuals in the United States except Massachusetts and Virginia due to those states’ restrictions. Patients on government insurance programs were also excluded. Samples of two tablets per medication lot were collected from mid-March 2020 to early August 2020 and one of each was analyzed in October 2020. Accepting two tablets was preferable to one to mitigate against the potential loss of one tablet. Participating individuals used Valisure’s sampling and shipping materials for sample collection and shipping. These sampling and shipping materials have been previously validated to contain no nitrosamine contaminants. The National Drug Codes (NDC) of metformin drug product samples were determined by visual inspection of unique tablet imprints and referencing the drugs.com database (see Supplemental Table 2).^39^ The cities and states where the samples were purchased were reported by those who volunteered samples for this study. Three combination products were accepted and included in dosage groups according to the metformin dose. The formulation as defined by the Abbreviated New Drug Application codes (ANDA) and current approval status for all samples was cross-referenced using the FDA Orange Book^40^ and National Drug Code (NDC) Directory.^41^

### Equipment, Supplies, and Chemicals

SCIEX EXIONLC AD high performance liquid chromatography coupled with SCIEX X500R time of flight high resolution mass spectrometry (HPLC-QToF HRMS) was purchased from SCIEX (Framingham, MA). InfinityLab Poroshell 120 EC-C18, 4.6 × 100 mm, 2.7 µm analytical column were purchased from Agilent (Agilent Technology, Santa Clara, CA). Certified reference material of NDMA, DMF, and isotope labeled DMF standard D_7_-DMF were purchased from Sigma-Aldrich (St. Louis, MO). Isotopic labeled NDMA standard ^13^C_2_-D_6_-NDMA was purchased from Cambridge Isotope Laboratories (Tewksbury, MA). All other chemicals and reagents were ACS or HPLC grade from Sigma-Aldrich.

### Sample Preparation

An entire single metformin tablet was used for sample preparation. One metformin tablet was weighed in a polypropylene tube and methanol was added at approximately 100 mg API/mL. The sample was homogenized, then centrifuged at 13,000 rpm for 1 minute. 250 µL of the supernatant was transferred and diluted with 750 µL of HPLC grade water, followed by vortex mixing. After filtering through 0.2 µm nylon filter, 480 µL of the sample extract was pipette-transferred to an amber LC vial, and 20 µL of isotopic internal standard mixture containing 20 ng of ^13^C_2_-D_6_-NDMA and 500 ng of D_7_-DMF is spiked into the sample extract. The final sample extract contains 40 ng/mL ^13^C_2_-D_6_-NDMA and 1 µg/mL D_7_-DMF, respectively. The vial is capped and vortexed prior to LC-HRMS analysis.

### Instrumental Analysis

NDMA in metformin was determined by LC-HRMS. Briefly, 10 µL of sample extract was injected, and chromatographic separation started at gradient of 98% of mobile phase A (0.1% formic acid in water) and 2% of mobile phase B (0.1% formic acid in methanol) at 0.6 mL/min and was held for 3.5 minutes. Mobile phase B ramped up to 98 % at 9.5 minutes and was held for 1.4 minutes. Then mobile phase B ramped down to 2% and was held to 15 minutes. NDMA elutes at 4.22 minutes, and DMF elutes at 4.53 minutes. Atmospheric pressure chemical ionization mode (APCI+) was selected to ionize NDMA and its isotopic labeled internal standard. Mass identification for NDMA was done by detecting the accurate mass of [M+1]^+^ in MRMHR acquisition mode for NDMA 75.0553, and ^13^C_2_-D_6_-NDMA 83.0997, respectively. The mass accuracy was set at 5 parts per million (ppm) and mass resolution for TOF MS/MS scan was greater than 31,000 for mass 176.09134 Da fragment of reserpine calibrant. Accurate masses of NMBA, NDEA, NDBA, NEIPA, and NDIPA were also monitored in the sample analysis.

### Quality Assurance and Quality Control

Quadratic calibration curve was established by a nine-point calibration ranging from 0.25 to 80 ng/mL for NDMA and 25 to 8000 ng/mL for DMF with both containing the same internal standard concentration as samples. Calibration is accepted if the r^2^ is equal to or greater than 0.99. The limit of quantification (LOQ) is defined as the lowest acceptable calibration point. The lowest calibration point must have a minimum signal to noise ratio of 10 in repeatability validation studies and calculated to be less than 20% different from its theoretical value. Concentrations of NDMA in samples were quantified by the internal standard method. Quality Control (QC) samples at low, medium, and high concentrations were analyzed by the same analytical procedure as metformin samples. The instrument LOQ was 0.25 ng/mL for NDMA and 25 ng/mL for DMF.

### Data Analysis

In order to compare concentrations of NDMA measured to the ADI limit, NDMA levels were scaled according to the common number of tablets taken per day at the US recommended maximum adult daily dose of 2,550 mg for IR metformin and 2000 mg for ER metformin (see Table 1).

**Table 1.** Information for crowdsourced samples including the number of immediate and extended release samples with NDMA concentrations that would result in exposure above the acceptable daily intake (ADI) at maximum tablet dose (2550 mg for IR, 2000 mg for ER).

| Metformin Unit Dose | Tablet Scaling Factor for Maximum Dose Per Day | Number of Immediate Release Samples | Number of Extended-Release Samples | Total |
| --- | --- | --- | --- | --- |
| 500 mg | 5.1 (IR) or 4 (ER) | 44 | 66 | 110 |
| 750 mg | 2.7 (ER) | NA | 13 | 13 |
| 850 mg | 3 (IR) | 1 | NA | 1 |
| 1000 mg | 2.6 (IR) or 2 (ER) | 30 | 1 | 31 |
| NDMA Above ADI | -- | 0% | 31% | 16% |

## Results

155 samples make up this data set (see Table 1) and participating individuals reportedly purchased the medication samples in 37 states.

Limit of quantification varied according to tablet weight but was between 5 to 12 ng per sample for NDMA and 500 to 1150 ng per sample for DMF. 49% of samples had detectable levels of NDMA and 74% had detectable levels of DMF. NDMA levels varied widely across samples (see Figure 1) while the majority of samples had high DMF concentrations. When scaled to the maximum adult tablet dosage equivalent, 16% of all samples had NDMA levels that exceed the acceptable daily intake. 31% of ER samples and none of the IR samples had NMDA concentrations that surpassed the ADI exposure limit at maximum tablet dose. Recovery efficiency for isotopically labelled NDMA was 92 ± 9%, and for isotopically labelled DMF was 96 ± 5%. No samples analyzed had detectable amounts of the nitrosamines NMBA, NDEA, NEIPA, NDIPA, or NDBA.

**Figure 1.**
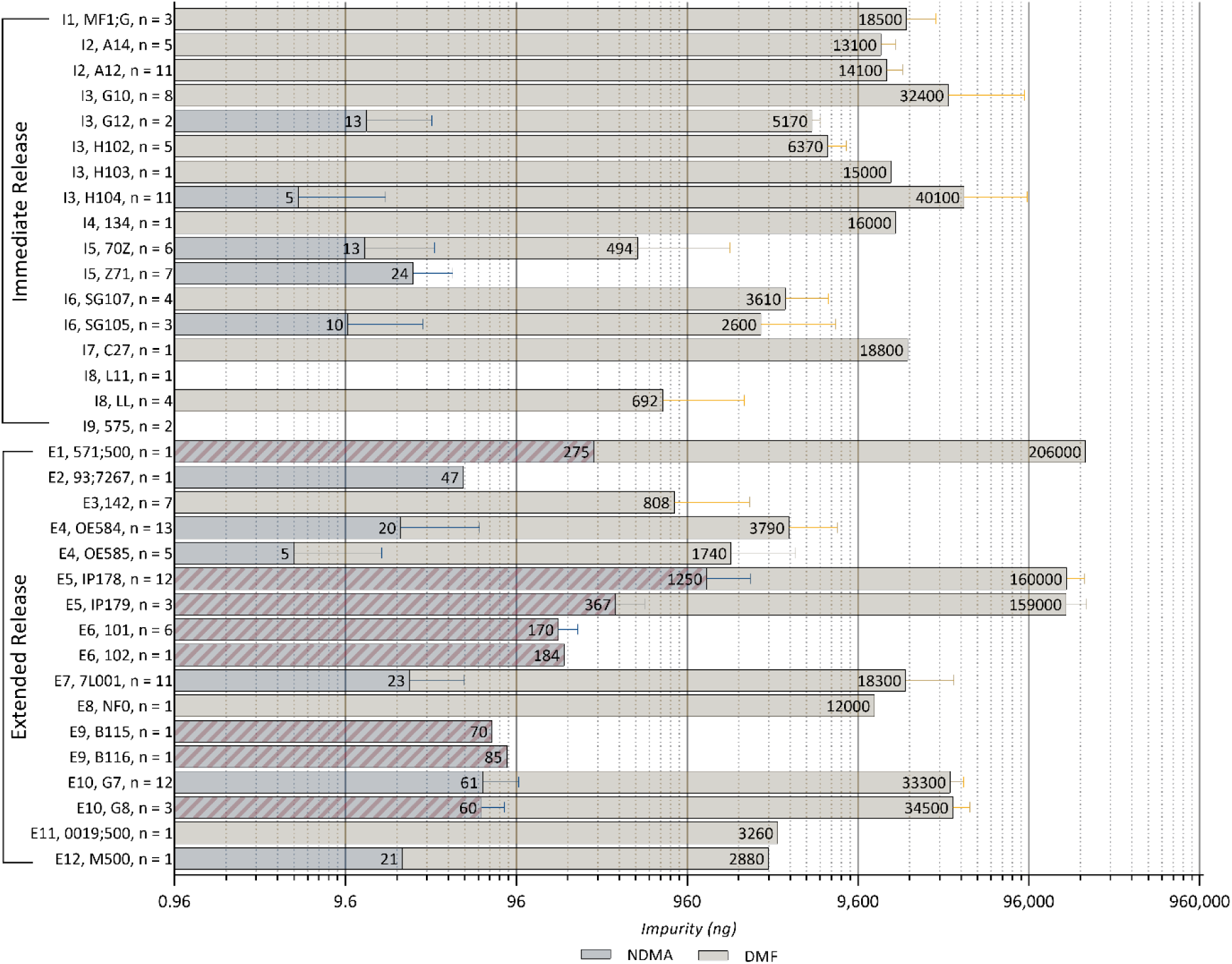
NDMA and DMF per maximum daily dosage of crowdsourced metformin samples, averaged by tablet imprint. Y-axis is labeled with formulation-ANDA code (see Supplemental Table 2), tablet imprint, and number of tablets with imprints sampled. Immediate and extended release formulations are indicated by I and E and marked, respectively. Imprints that correspond to products that have been recalled for NDMA in 2020 are indicated with striped NDMA data bars. Error bars represent one standard error of the mean (SEM).

Comparison of IR and ER formulations yielded distinct results (see Figure 2). Of the 155 tablets sampled, 80 (52%) were ER and 75 (48%) were IR. 58 (73%) and 59 (74%) of ER samples had detected levels of NDMA and DMF, respectively. 48 out of the 80 ER tested (60%) had both compounds present, 11 (14%) had neither, 11 (14%) had DMF but no NDMA, and 10 (13%) had NDMA but no DMF. 25 ER samples had concentrations of NDMA that would exceed the NDMA ADI exposure limit if taken at maximum daily dosage. The ER sample containing the greatest quantity of NDMA when scaled, a tablet with imprint IP178, was equivalent to 2,993 ng, or 31 times the ADI. A sample of a tablet with the same imprint also had the highest quantity of DMF when scaled, equivalent to 225 µg. Out of the 12 tablets sampled with this particular imprint, the lowest measurements of the two compounds were 96 ng NDMA and 76 µg of DMF, which were measured in the same sample.

**Figure 2.**
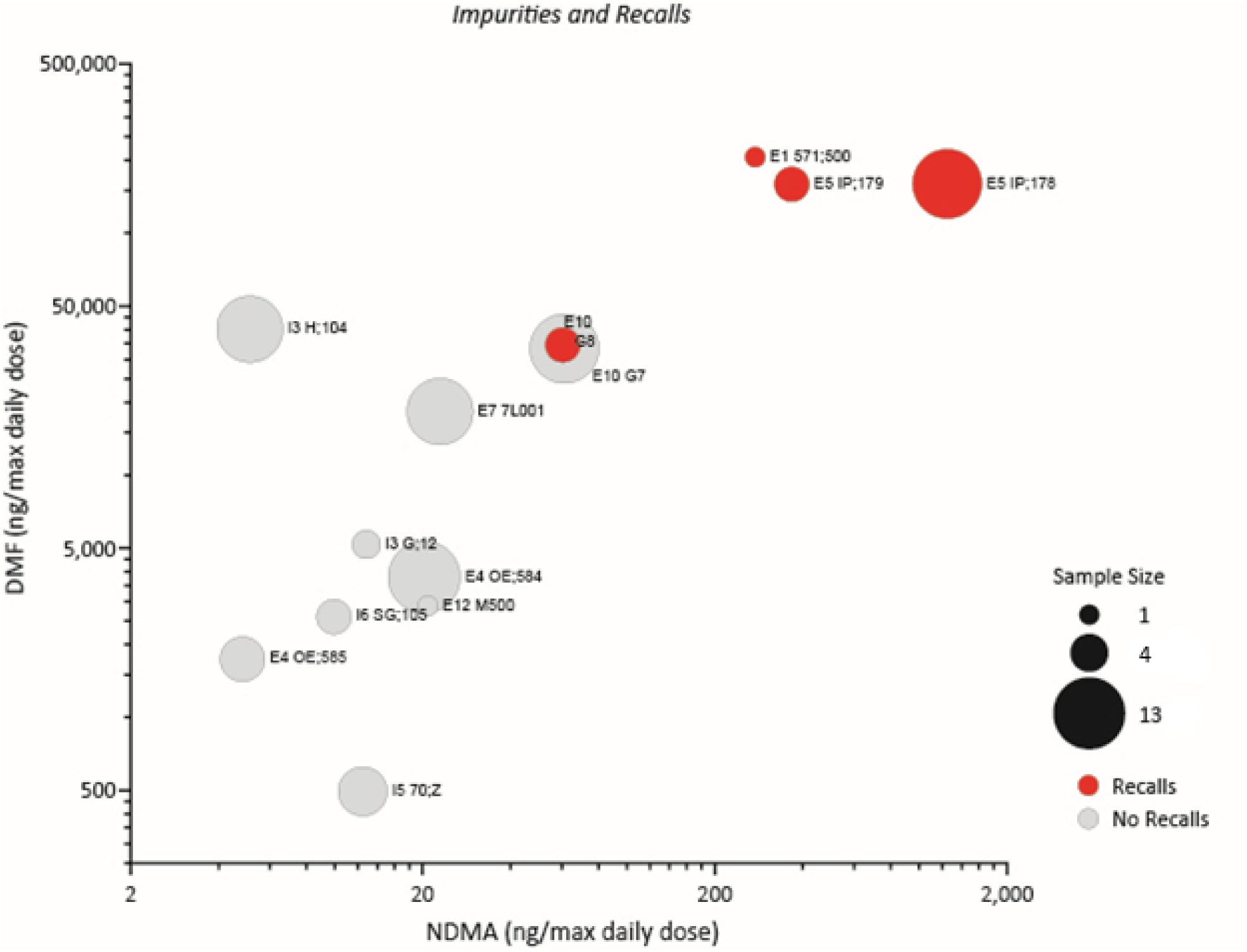
DMF versus NDMA detection in nanograms (ng) per maximum daily dosage of crowdsourced metformin samples, averaged by tablet imprint. Bubble size indicates number of tablets of the same imprint sampled and are labeled with ANDA code and tablet imprint. Axes are on a log scale, therefore, only imprints for which both NDMA and DMF averages were measured to be above zero are presented here. Imprints that correspond to NDCs that have been recalled for NDMA^43^ are colored red.

IR tablet formulations yielded results suggesting lower overall levels of contamination than ER tablets. 18 (24%) and 55 (73%) samples of IR had detected levels of NDMA and DMF, respectively. 10 out of the 75 tested (13%) had both, 12 (16%) had neither, 45 (60%) had DMF but no NDMA, and 8 (11%) had NDMA but no DMF. None of the IR formulations exceeded 96 ng NDMA when scaled to max daily dosage. The highest quanitity of NDMA detected in an IR tablet when scaled was equivalent to 40 ng. 6 samples with this imprint were tested and only 2 of them had results which exceeded the LOQ. The highest quantity of DMF detected in an IR tablet when scaled was equivalent to 173 µg. Of the 8 samples with this imprint tested, the lowest contained 3 µg of DMF.

## Discussion and Conclusions

Quantitative NDMA testing reported here for 155 crowdsourced metformin drug products suggest strong correlation between drug quality and specific National Drug Codes (NDC), which are registered to the company that labels the product for sale in the United States. Of greatest concern are labeling companies with NDMA concentrations that could result in NDMA exposure above the acceptable daily intake (ADI) limit established by the FDA. The exposure to NDMA is compounded by the fact that metformin often treats chronic diseases, and as a result, the cumulative exposure to this contamination for many patients may have occurred for many years. Additional exposure to DMF contamination may exacerbate this problem. Interestingly, DMF was found at high levels in both IR and ER formulations while NDMA was found less frequently in IR samples. It is plausible that addition of different inactive ingredients could potentiate formation of NDMA in ER compared to IR, whether from degradation of DMF or by another mechanism. Although there appear to be some trends for specific ANDAs and labeling companies, the overall dispersed nature of contamination suggests complexity in the source of the NDMA and DMF.

The results reported here are derived from sampling of a limited cross section of crowdsourced on-market products. Importantly, medications purchased by the government (e.g., Department of Defense, Veterans Health Administration) and dispensed by federal pharmacies may have different purchasing agreements for metformin compared to civilian pharmacies. A phone survey to four federal pharmacies to inquire about the labelers of their stocks of metformin at the time of sampling revealed five companies contained in this study and one not represented in this study (data not shown). Therefore, it is plausible that individuals receiving medications from government sources experience similar contamination issues, but further research is necessary.

Single tablet analysis employed in this study does not provide statistical significance for nitrosamine content results at the lot or batch-level or for some formulations. In some cases, multiple medications with the same ANDA as determined by tablet characteristics (see Supplemental Table 2) gave similar results but further surveillance is needed to determine whether trends are significant. Only two tablets were requested from individuals to minimize impact on patients’ medication regimen. Importantly, 16% of samples had NDMA levels that exceeded the ADI limit at maximum dosage per day. This finding is consistent with data from Valisure’s FDA Citizen Petition on Metformin^29^ reporting 20% for the same metric for lots tested in triplicate using methodology that separates NDMA from DMF (Note, Valisure’s citizen petition cited 42%, which was an overestimate due to high DMF concentration interference in the original method used). Notably, Valisure’s FDA Citizen Petition predated all metformin recalls for NDMA in the US while a portion of this study’s sampling timeframe included broad, all-lot recalls from multiple manufacturers; therefore, this study should reflect lower NDMA detection as an outcome of successful recalls. Interestingly, fewer instances of NDMA detected above the ADI limit per tablet or when scaled to maximum daily dose occurred for IR formulations compared to ER formulations (0% and 31% respectively, see Table 1).

The distribution of the limited tested products in this study is unable to fully predict the prevalence of product formulations or NDCs that are on market in the US or their stability. Rarer drug products containing metformin were predictably absent from this study. For example, 750 mg ER drug products were submitted by few individuals and only one 850 mg IR metformin drug product was submitted. Likewise, only three combination products that included metformin were submitted. Less commonly prescribed metformin drug products are of interest since they are likely not manufactured as ubiquitously as 500 and 1000 mg IR and ER products. Despite limitations in sampling, products measured with high NDMA are linked by NDC to products that have been recalled for NDMA suggesting agreement with industry testing (see Figure 2). Recently, FDA announced voluntary recalls of Metformin ER 750 mg due to NDMA detection at 17-months stability study.^42^ This may suggest that NDMA can be formed at storage conditions even if not presented in API or finished drug product when released. In the interest of public health, results presented here support the need for further study of metformin drug product stability in distribution and storage environments.

Despite sampling limitations, this novel approach to crowdsourced medication samples for contamination analysis may represent the least biased snapshot of post-market drug product quality. The broad distribution of NDMA and DMF contamination levels across metformin products warrants further investigation given the possibility for long-term exposure as a maintenance medication. If the distribution of contamination observed in this study is indicative of the US market and has been consistent over time, then in 2019^4^ approximately 13.6 million prescriptions were unacceptably contaminated with NDMA and approximately 62.9 million prescriptions exposed patients to DMF. The highest amount of NDMA detected in this study was 748 ng in a single 500 mg dosage unit (1.5 parts per million API). If four of these tablets are taken per day, as is commonly prescribed, the NDMA exposure would be 31-times greater than the ADI limit established by the FDA. For the NDCs listed in this study showing high levels of NDMA and DMF, further investigation and product recalls are warranted to protect the American public from unnecessary exposure to these probable carcinogens.

## Data Availability

Granular data is available in the supplemental table and further requests may be made to the corresponding author.

---

**Supplemental Table 2.**
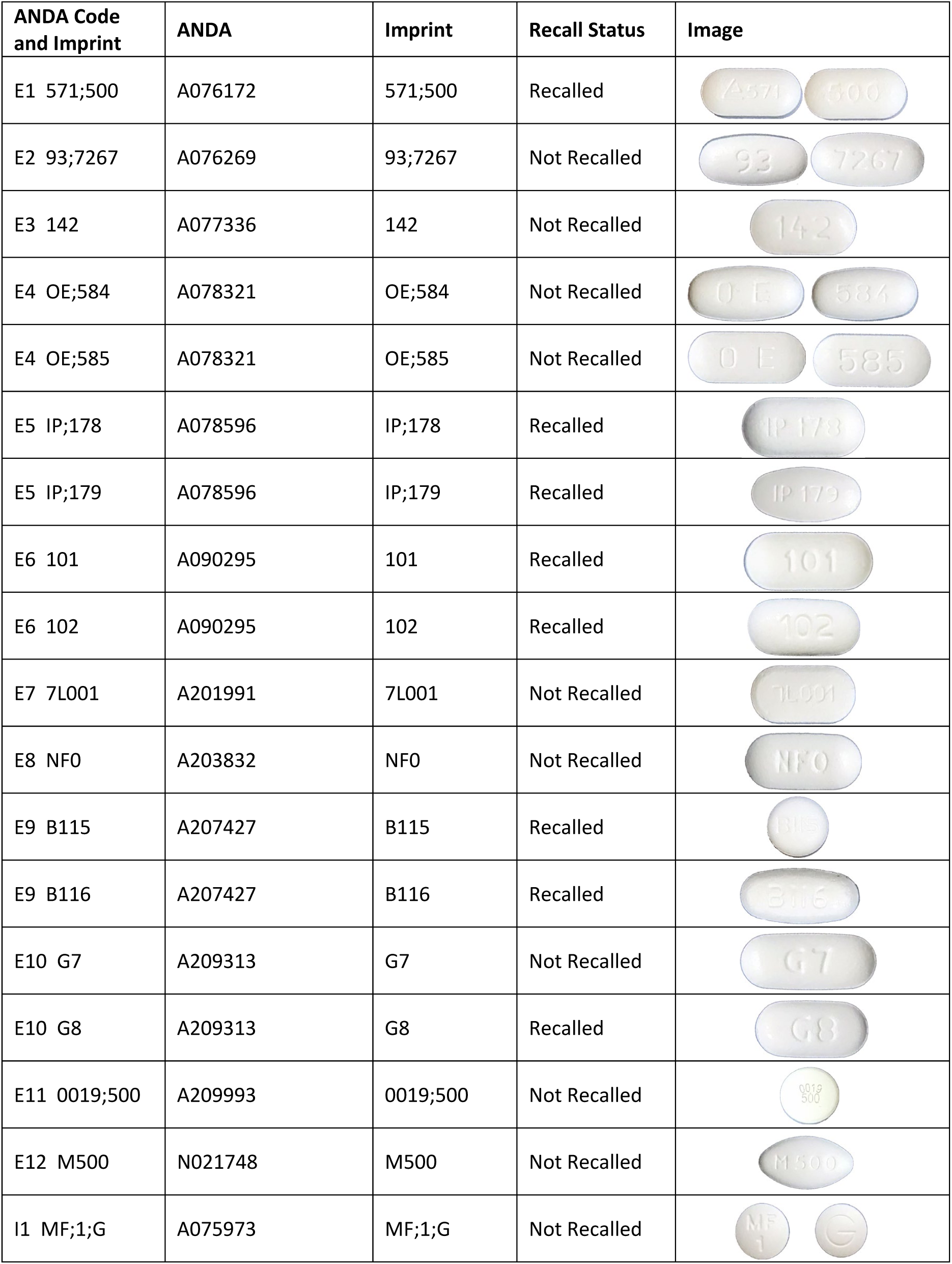
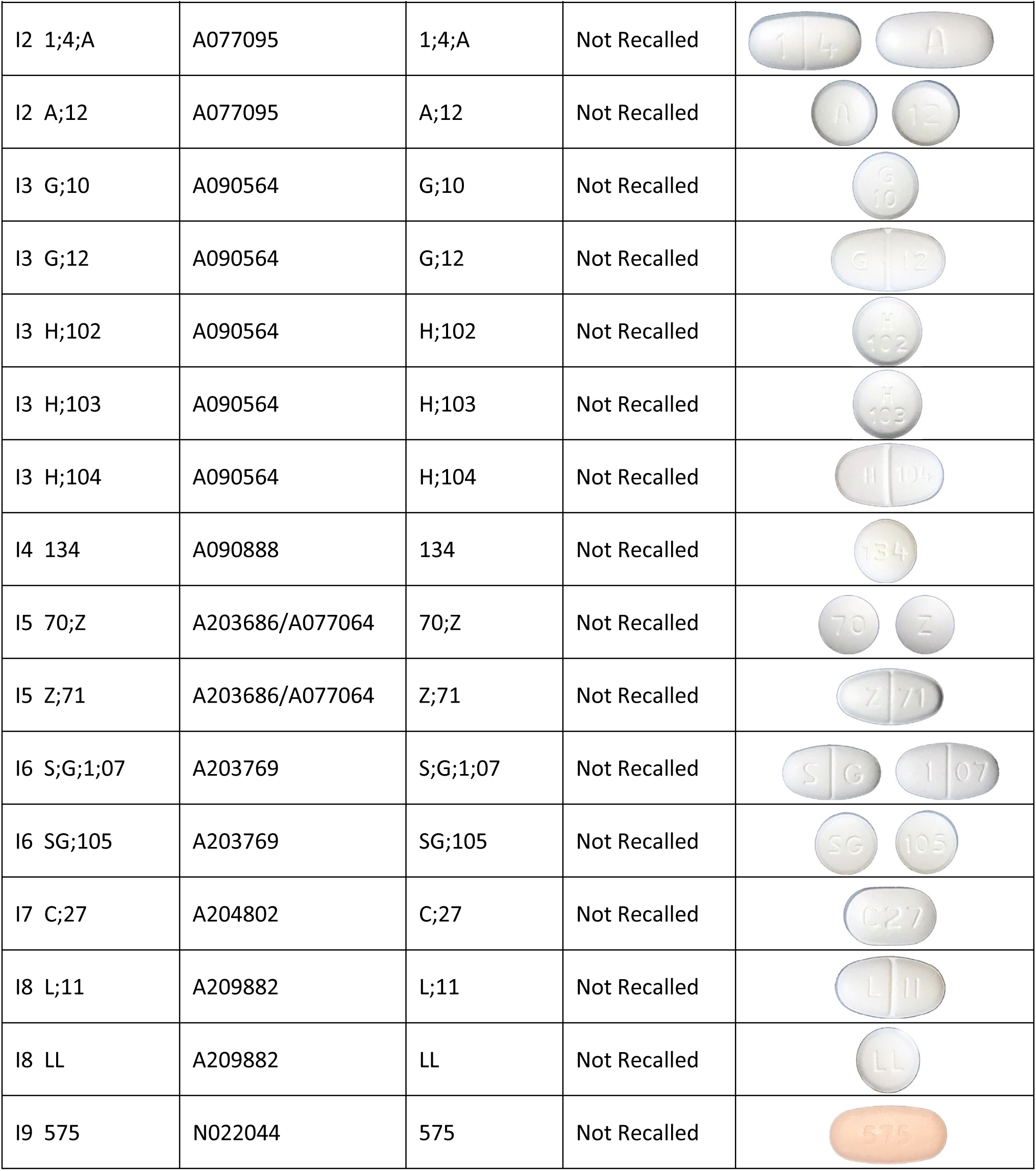
Additional information for crowdsourced samples including the figure codes and ANDA determined from tablet imprints. 2020 recall status for metformin drug products that share an ANDA with study samples are indicated.

| ANDA Code and Imprint | ANDA | Imprint | Recall Status | Image |
| --- | --- | --- | --- | --- |
| E1 571;500 | A076172 | 571;500 | Recalled |  |
| E2 93;7267 | A076269 | 93;7267 | Not Recalled |  |
| E3 142 | A077336 | 142 | Not Recalled |  |
| E4 OE;584 | A078321 | OE;584 | Not Recalled |  |
| E4 OE;585 | A078321 | OE;585 | Not Recalled |  |
| E5 IP;178 | A078596 | IP;178 | Recalled |  |
| E5 IP;179 | A078596 | IP;179 | Recalled |  |
| E6 101 | A090295 | 101 | Recalled |  |
| E6 102 | A090295 | 102 | Recalled |  |
| E7 7L001 | A201991 | 7L001 | Not Recalled |  |
| E8 NF0 | A203832 | NF0 | Not Recalled |  |
| E9 B115 | A207427 | B115 | Recalled |  |
| E9 B116 | A207427 | B116 | Recalled |  |
| E10 G7 | A209313 | G7 | Not Recalled |  |
| E10 G8 | A209313 | G8 | Recalled |  |
| E11 0019;500 | A209993 | 0019;500 | Not Recalled |  |
| E12 M500 | N021748 | M500 | Not Recalled |  |
| I1 MF;1;G | A075973 | MF;1;G | Not Recalled |  |
| I2 1;4;A | A077095 | 1;4;A | Not Recalled |  |
| I2 A;12 | A077095 | A;12 | Not Recalled |  |
| I3 G;10 | A090564 | G;10 | Not Recalled |  |
| I3 G;12 | A090564 | G;12 | Not Recalled |  |
| I3 H;102 | A090564 | H;102 | Not Recalled |  |
| I3 H;103 | A090564 | H;103 | Not Recalled |  |
| I3 H;104 | A090564 | H;104 | Not Recalled |  |
| I4 134 | A090888 | 134 | Not Recalled |  |
| I5 70;Z | A203686/A077064 | 70;Z | Not Recalled |  |
| I5 Z;71 | A203686/A077064 | Z;71 | Not Recalled |  |
| I6 S;G;1;07 | A203769 | S;G;1;07 | Not Recalled |  |
| I6 SG;105 | A203769 | SG;105 | Not Recalled |  |
| I7 C;27 | A204802 | C;27 | Not Recalled |  |
| I8 L;11 | A209882 | L;11 | Not Recalled |  |
| I8 LL | A209882 | LL | Not Recalled |  |
| I9 575 | N022044 | 575 | Not Recalled |  |

